# Climate blind spots in malaria control: Frontline perspectives on health system readiness in Zambia

**DOI:** 10.64898/2025.12.05.25341687

**Authors:** Nyuma Mbewe, Therese Sherma Nzaisenga, Kelvin Mwangilwa, Jonathan Mwanza, Stephen Bwalya, Ignitius Banda, Cheepa Habeenzeu, Paul Zulu, Loveness Nikisi, Nathan Kapata, Allan Mayaba Mwiinde

## Abstract

**Background:** Climate change is increasingly recognised as a significant barrier to malaria elimination, especially in low-and middle-income countries (LMICs), where vulnerability to vector-and waterborne diseases is heightened. Climate variability increasingly influences malaria transmission dynamics, yet its impact on malaria control efforts remains underexplored. This study explored healthcare workers’ and community-based volunteers’ (CBVs) perspectives on climate change and the perceived contribution of climate variability to malaria transmission in Zambia.

**Methods:** A cross-sectional qualitative study was conducted between August and October 2023 across twenty purposefully selected districts representing high-and low-burden malaria settings. Nine key informant interviews and fourteen focus group discussions were conducted with malaria program officers, clinicians, environmental health officers and CBVs. Data were transcribed verbatim, imported into ATLAS.ti version 23, and analysed thematically.

**Results:** Participants consistently reported that flooding, drought, deforestation, and shifting rainfall patterns were increasing mosquito breeding sites and altering malaria transmission seasons. Climate-related disruptions, poor road access during floods and competing health priorities, including cholera outbreaks and COVID-19, were perceived to hinder effective malaria prevention and case management. While participants acknowledged the need for a more integrated response, they largely emphasised reinforcing existing malaria control strategies, such as indoor residual spraying (IRS) and insecticide-treated nets (ITNs), with limited reference to broader climate adaptation measures or national climate policies, highlighting gaps in policy dissemination and implementation. Participants also noted contextual barriers, including vector resistance and diagnostic inaccuracies. Notably, the emerging role of malaria vaccination was not mentioned, indicating a potential knowledge gap in climate-adaptive malaria strategies.

**Conclusions:** Frontline perspectives highlight substantial climate-related challenges to sustaining malaria control in Zambia and gaps in climate-health knowledge among HCWs and CBVs. Strengthening climate-resilient systems, improving policy dissemination and integrating climate adaptation into malaria programming and training are critical to sustaining progress towards elimination.

**Author Summary:** Despite clear evidence that climate change is reshaping malaria transmission in sub-Saharan Africa, little is known about how frontline health workers perceive and respond to these shifts. This study provides the first multi-district qualitative examination of healthcare worker and community volunteer perspectives on climate–malaria interactions in Zambia. Our findings reveal critical knowledge gaps, limited awareness of existing climate–health policies, and an over-reliance on traditional malaria interventions that fail to integrate climate-resilient strategies. These insights underscore a pressing need for targeted training, strengthened policy dissemination, and multisectoral collaboration to build climate-ready malaria programmes. By illuminating the disconnect between climate science and frontline practice, this study highlights a fundamental barrier to sustaining malaria elimination in a rapidly changing climate.

## Introduction

The World Health Organization (WHO) recognises climate change as the biggest health threat to humanity (1,2). Its impact is more pronounced in low-and middle-income countries (LMICs), where vulnerability to vector-and waterborne diseases is heightened (3). Malaria transmission seasons are expected to lengthen and intensify due to climate change (3–5). These effects will be compounded by population growth, malnutrition from crop failures, worsening existing health challenges, and a fragile public health infrastructure.

Malaria remains one of the world’s deadliest diseases, with cases and deaths continuing to rise despite billions invested in control strategies, hindering the 2030 Malaria Elimination Agenda (6,7). In 2023, there were over 263 million cases and 597,000 deaths, up from 252 million cases in 2022(8). Sub-Saharan Africa (SSA) accounted for 94% of cases and 95% of all deaths globally. WHO now recommends tailored, data-driven approaches, including indoor residual spraying (IRS), insecticide-treated nets (ITNs), chemoprophylaxis, enhanced surveillance, and malaria vaccination for children under five (7,9). More than 30 endemic SSA countries, including Zambia, have adopted these interventions (9).

Climate change is a major barrier to malaria elimination (6,9). Predictions modelled around a worst-case scenario climatic pattern suggest an additional 75.9 million people at risk from endemic (10-12 months) exposure to malaria transmission in Eastern and Southern Africa by the year 2080, with the greatest population at risk in Eastern Africa (10). Temperature variations have been linked to changes in malaria incidence rates in SSA, with lower incidence observed following decreases in temperature and, conversely, rising temperatures associated with rising incidence (11). Globally, rising temperatures and shifting rainfall patterns have increased the suitability of conditions for malaria transmission by 40% (9). Favourable conditions for Anopheles mosquitoes and El Niño’s role in malaria incidence further stall progress (3). Environmental factors such as humidity and aridity also influence malaria transmission, with only Algeria, Egypt, and Cape Verde certified as malaria-free by the WHO (12).

Despite the revised Malaria Elimination Strategy (2021) in Zambia, malaria mortality increased by 3.2% from 2020 to 2022 (12). Transmission peaks between January and April, aligning with the rainy season (9,13). Shamponda et al. found a strong correlation between rainfall and malaria incidence, with humidity, land use changes, vapour pressure, and temperature also playing a role (14). Additionally, the seasonal variability of temperature fluctuations has been seen to be directly linked with fluctuations in several SSA countries, including Zambia (10,15).

WHO recommends climate-resilient and climate-informed healthcare systems to support high-quality care and universal health coverage (16). A well-trained and protected health workforce is essential, yet many clinicians and non-clinicians lack training on climate change (15). While healthcare workers acknowledge climate change’s impact on health, knowledge gaps remain regarding mitigation strategies (17). These gaps risk undermining malaria elimination efforts, particularly as climate-driven shifts in transmission are becoming more pronounced.

This study forms part of a larger mixed-methods analysis examining climate change’s influence on malaria transmission in Zambia over 15 years. In this qualitative component, we explore healthcare workers’ and community-based volunteers’ (CBV) perceptions of how climate change affects malaria interventions and health system readiness, while the quantitative findings are reported separately. Understanding these trends will inform climate-adaptive strategies for malaria control and strengthen Zambia’s health system resilience.

## Materials and Methods

### Study site

This study was conducted in Zambia, a landlocked sub-Saharan African country with a surface area of 752,612 km² and a population of 19.6 million (2022 Census) (18). It is divided into 10 provinces and 116 districts. Zambia has a tropical climate, with higher rainfall in the north. The rainy season peaks from October to April, but recent years have seen increased flooding due to El Niño, followed by droughts (19). Malaria is endemic and perennial, with Lusaka and Southern provinces recording the fewest cases annually (20). Plasmodium falciparum, transmitted by female Anopheles mosquitoes, causes 90% of cases and is known to be sensitive to climate variables (21).

### Study design

This study constitutes the qualitative component of a mixed-methods cross-sectional study assessing the effects of climate variability factors, such as rainfall patterns, humidity, and temperature, on malaria incidence in Zambia. For this qualitative arm, we focused specifically on understanding frontline perceptions of climate-malaria interactions and health system readiness. Purposive sampling was conducted for the districts with the highest and lowest malaria incidence in each province, based on the previous year’s Health Information Management System (HMIS) data.

Qualitative data were collected from August to October 2023, focusing on healthcare workers’ perspectives on climate change’s impact and mitigation strategies. Key Informant Interviews (KII) and Focus Group Discussions (FGD) were held at two district hospitals per province (Fig 1). This design enabled exploration of both individual expertise and shared team experiences in malaria control within a changing climate.

**Figure 1:**
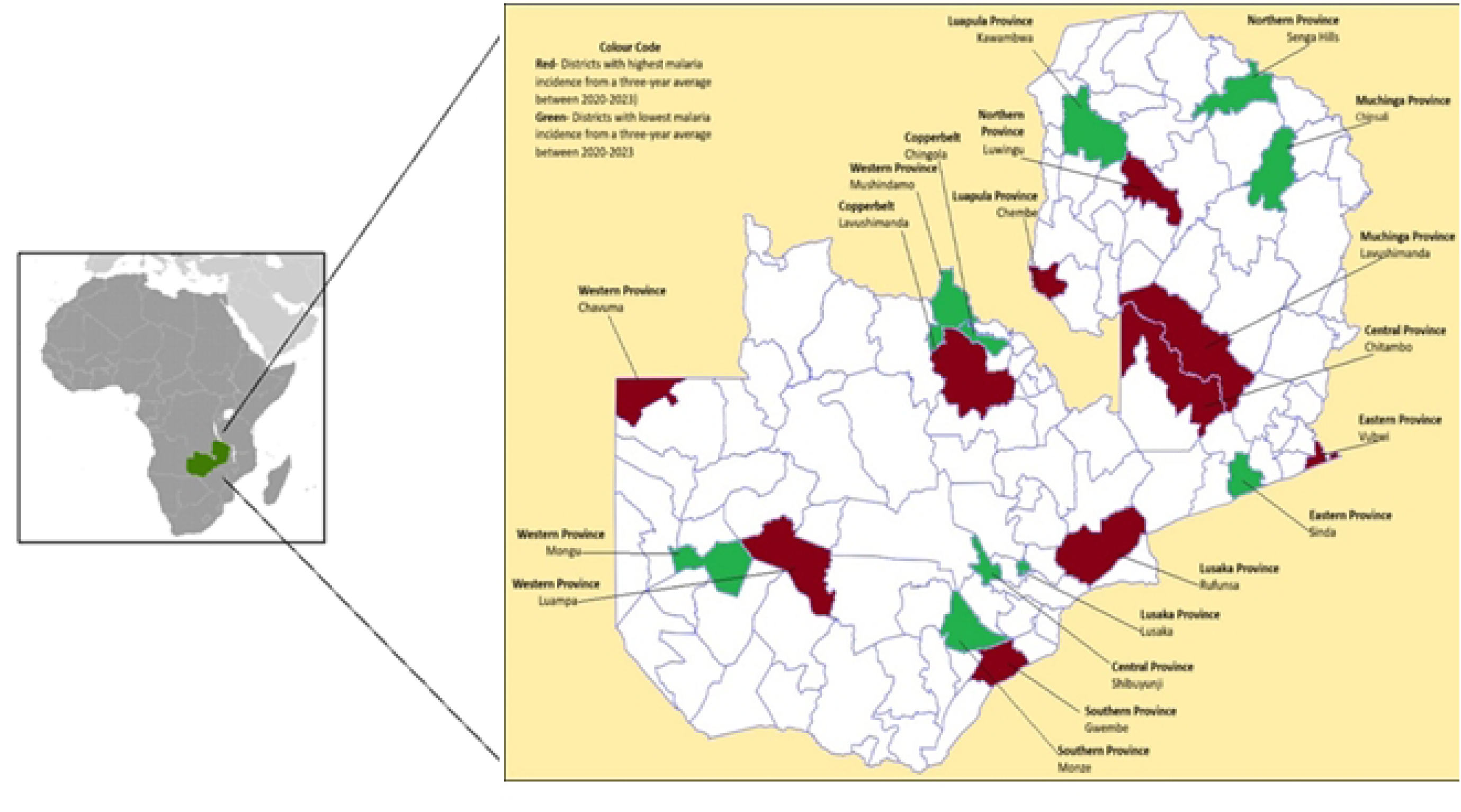
Study Sites for Key Informant Interviews and Focus Group Discussions. Map of Zambia showing the 20 purposively selected districts where qualitative interviews (KIIs and FGDs) were conducted to explore healthcare workers’ and community-based volunteers’ perspectives on climate change and malaria control. Districts represent high-incidence and low-incidence settings across all 10 provinces.

### Study population

To capture perspectives across the health system, data were collected at the facility or district levels. Purposive sampling targeted participants with over three years in malaria control or involvement in climate change programs. Key Informant Interviews (KII) were conducted with District Health Directors, Public Health Specialists or Malaria Program Officers. At each facility, the Malaria Program Officer, usually an Environmental Health Officer (EHO), was interviewed.

KIIs lasted approximately 45 minutes in person. FDGs with four to seven malaria programme staff, including CBVs, clinicians and nurses, lasted about 90 minutes to ensure full participation. This mix of cadres ensured representation from both technical and community-facing components of malaria control.

### Sampling and data collection procedure

Data for this study and its companion quantitative study were collected across all 10 provinces. A detailed facility selection process is available in the supplementary materials. In total, 20 districts were selected (Fig 1). KII used structured, open-ended guides, allowing emerging themes to shape subsequent interviews. Interviews were conducted in English. After nine KII and fourteen FDGs, thematic saturation was achieved, and no new concepts emerged (Table 1). The questionnaire is available on request.

**Table 1.** Distribution of participants across cadres and levels of the Health System.

| Type of Cadre | Primary Healthcare Facility workers (n=38) | District Office bearers (n=35) |
| --- | --- | --- |
| Community-Based Volunteer (CBVs) | 29 | — |
| Environmental Health Officers/Technicians (EHOs) | — | 17 |
| Clinical Officer General (COG) | — | — |
| Nurses | — | 7 |
| Medical Doctors | — | 2 |
| Laboratory Technicians | — | 1 |
| Others (Pharmacy, Nutritionist, Surveillance officers, Monitoring and Evaluation, District Health Information Officer) | — | 8 |
| **Total** | **38** | **35** |

### Data analysis

Data captured using digital recorders during KII and FGDs were transcribed verbatim by trained transcribers, including nonverbal cues. Transcripts were proofread against the audio to ensure accuracy. The participants’ names were omitted.

Investigators (NM and JM) audited transcripts by cross-checking with recordings to preserve meaning. Minor edits improved readability, correcting misheard words, vernacular phrasing, and grammar.

Qualitative analysis began with repeated transcript review to identify themes related to malaria incidence and climate change. Themes were developed both deductively (from existing theories) and inductively (emerging from data). Key themes included malaria incidence trends, intervention successes and challenges, climate change knowledge, its impact on malaria, and possible mitigation strategies. A codebook was created to define themes and guide further analysis as previously described (22).

Transcripts were imported into ATLAS.ti version 23 for review. Each transcript was analysed line by line, noting relevant statements. The interviewers’ summary notes were also reviewed to capture nonverbal cues not evident in recordings. The final thematic structure reflects patterns consistently observed across the provinces, including increasing malaria trends and influencing factors such as reduced funding and the COVID-19 pandemic. Results are presented in narrative form, supported by direct participant quotes.

### Ethical Considerations

Ethical approval was obtained from the Lusaka Apex Medical University IRB and the National Health Research Authority (Ref No: NHRA000013/23/08/2023). Provincial Health Directors were notified about the study, especially its qualitative aspects. Interviewees provided written informed consent, and all interviews were anonymized.

## Results

### Context within which malaria control strategies are delivered

A total of 73 individuals participated in the interviews. 17.94% were aged 20-29, 41.03% were 30-39, 20.51% were 40-49, and 20.51% were 50 or older. Participants had served in their roles for three to seventeen years. All study facilities were part of the national malaria control program. The District Health Office, led by the District Health Director, oversees health facilities and public health services. Malaria program officers, often environmental health or surveillance officers, handle supervision, monitoring, training, and coordination. Clinical officers, nurses, and doctors managed patient care, while CBVs supported Integrated Community Case Management (ICCM) for early diagnosis and treatment.

CBVs also played a key role in community education and intervention delivery, yet lacked formal training on climate-health interactions. District program officers meet quarterly to review progress and challenges. Table 1 summarizes participant professions, with EHOs and CBVs making up the largest group.

### Factors associated with knowledge about climate change

All respondents were knowledgeable about malaria control, given their roles in patient management or the national malaria program. All FGD participants (100%) reported awareness of climate change, though the depth of understanding varied widely.

Importantly, awareness of climate change did not necessarily translate into understanding of climate-malaria links or health system adaptation needs, reflecting a climate knowledge-practice gap.

Self-reported perceptions of climate change’s impact on malaria incidence are summarised in Table 2 and further explored in the narrative.

**Table 2:**
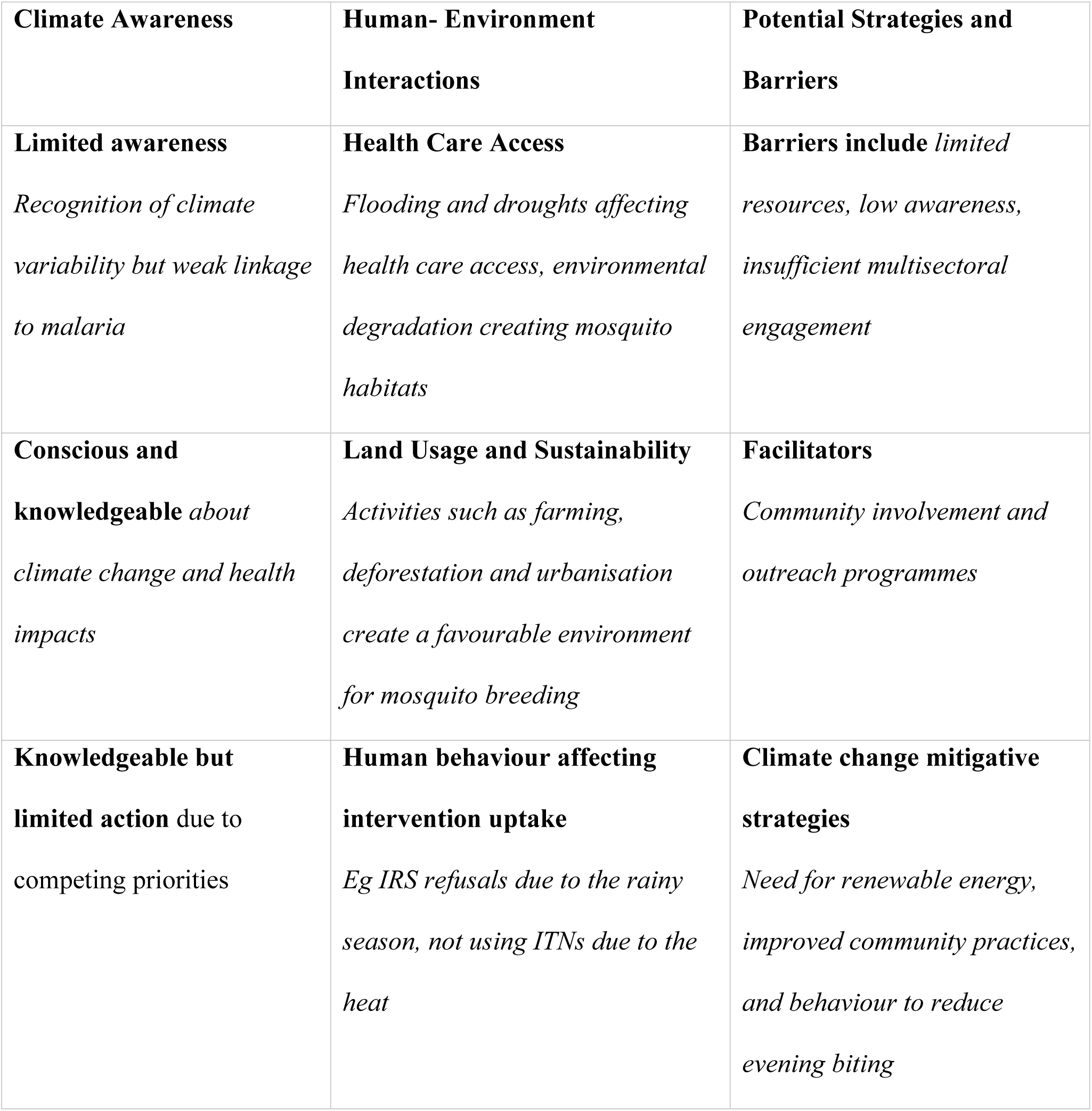
Thematic Overview of Climate Awareness, Human-Environment Interactions and Potential Strategies.

### Awareness of the detrimental effects of climate change

Healthcare workers had varying awareness of climate change’s impact on malaria and other infectious diseases. The study highlighted gaps in understanding of how specific climatic factors, such as humidity, rainfall variability and heat, drive malaria transmission, limiting the development of appropriate mitigation or adaptation strategies.

Some participants linked climate change to human-environment interactions that could increase malaria endemicity, as illustrated by these quotes from district malaria control officers.

> *“Malaria incidence is strongly influenced by climate. For instance, if the rains begin earlier in October, we see a rise in cases starting from then rather than in December. Additionally, during hot periods, we experience a surge in diarrheal cases because as the water table drops, people start drawing water from streams and shallow wells, which also serve as mosquito breeding sites.” – Male Malaria Program Officer, FGD3*.Despite this, most participants were unaware of national climate adaptation policies or the need for climate-resilient malaria strategies, reinforcing the “climate blind spots” seen across districts.

Participants’ self-ratings of climate change knowledge varied independently of training, education level or years of service. In one KII the respondent to the question “How much do you know about this topic of climate change?” affirms *“Very much. I did my bachelor’s in environmental studies.”* (*Male EHO in FDG7)* whilst in a different FGD the respondent acknowledged “*I do not know much, I will rate myself at 30%.”-Female EHO, FGD 3*.

### Observed influence of climate change on health

The healthcare workforce reported a U-shaped impact, with increased malaria cases during both flooding and droughts. Participants described challenges such as misaligned timing of indoor residual spraying (IRS), migration to fishing or farming camps after IRS and reduced access to antenatal IPT during floods. These perceptions reflected real-world implementation barriers rather than climate-adaptive service planning:

> *“IRS isn’t very effective due to delayed applications during the rains. Mushindamo also has many impassable areas because of poor road networks, so not everyone is reached during ITN distribution. Additionally, during floods, pregnant women can’t visit the clinic for IPT.” – Male EHT, FGD 5*.Other participants linked climate change to expanding mosquito breeding sites and to indirect pathways, such as food insecurity, which contributes to anaemia and severe malaria.

> *“Recently, there’s been a greater need for spraying interventions due to the increase in breeding sites around the district.” – Male EHT, KII2*.

### Contextual factors influencing the effectiveness of malaria control strategies

Respondents identified multiple contextual stressors that undermine malaria control, including the COVID-19 pandemic, supply chain constraints, and concurrent outbreaks such as cholera. These competing priorities diverted attention and resources from malaria and revealed the fragility of health systems’ ability to absorb shocks, exacerbating climate-related vulnerabilities. New interventions for climate change mitigation and adaptation should integrate with existing programs to build health system resilience and avoid overwhelming the system or duplicating efforts.

> *“As a surveillance officer, we didn’t focus much on malaria during COVID because we were overwhelmed.” – Male EHT, FGD 1*.Workers emphasised the need for climate change interventions that integrate with existing programmes rather than overwhelm them.

### Proposed mitigation and adaptation measures

Respondents identified human-environment interactions, such as farming, deforestation, and urbanisation, as key contributors to both climate change and increased malaria transmission. They recommended community-based measures such as reforestation and conservation farming, improved drainage, and the promotion of green energy.

> *“In rural areas, people continue cutting trees as it’s their main source of income, especially for charcoal production. Without electricity, they have no alternatives for cooking and survival.” – Male COG, FGD 2*.However, most participants focused on strengthening existing malaria tools (IRS, ITNs, larviciding) rather than proposing climate-resilient strategies for health facilities, workforce capacity or early warning systems, underscoring a system-wide adaptation gap.

> *“It’s our job as community workers to help sensitize the community on climate change and malaria.” – Female CBV, FGD 6*.

> *“Education alone won’t be enough; we need ITNs, repellents, larviciding, and mass anti-malaria drug administration twice a year.” – Male EHT, KII 4*

Notably, no respondents mentioned the role of malaria vaccines as a climate-resilient strategy, nor did they suggest strengthening facility climate readiness, e.g., heat-adapted wards and resilient supply chains.

This reveals a significant climate-awareness gap even among experienced malaria personnel, reinforcing the study’s primary finding – the absence of climate-informed health system planning.

## Discussion

This study explored how healthcare workers (HCWs) and community-based volunteers (CBVs) understand climate change and its implications for malaria control, and how these perceptions can shape proposed mitigation and adaptation strategies. This qualitative component provides critical context for interpreting stalled progress in malaria elimination and complements the quantitative findings reported separately.

Consistent with global assessments, participants recognised that climate variability is increasingly influencing malaria dynamics in Zambia (23). They demonstrated strong knowledge of core malaria countermeasures but less clarity and confidence in explaining the mechanisms through which climate change affects malaria transmission. This mirrors gaps that are identified in the Zambia National Health Climate Adaptation Plan (NHCAP) and similar studies across sub-Saharan Africa, where knowledge of climate-health linkages is uneven despite high frontline exposure (14,19,24). These findings are similar to assessments done elsewhere in the world (17,25).

Importantly, HCWs attributed recent reductions in malaria burden largely to decentralised diagnosis and treatment, particularly the expanded role of CBVs in ICCM, aligning with the National Malaria Elimination Strategic Plan (NMESP) 2022-2026 (13). However, participants also reported that malaria incidence has plateaued, attributing this partly to behavioural and environmental barriers such as net non-use during hot seasons, deforestation and reliance on shallow wells for household water use (14,26). Other contributing factors noted in the climate-health matrix concerning malaria included the rising unpredictability of climate-related events, such as early rains, extreme heat, flooding, and prolonged droughts (13,27). These observations are consistent with ecological system analyses from the region, which show the impact of land-use changes, humidity, and rainfall patterns on vector breeding (11,28,29).

While respondents articulated an awareness of climate variability, their understanding was not routinely translated into climate-resilient malaria strategies. Most HCWs emphasised reinforcing traditional interventions such as IRS, ITNs and case management, while under-emphasising adaptation strategies such as improving facility climate resilience, adopting renewable energy sources, climate-proofing supply chains or integrating surveillance with meteorological data. Few referenced national climate adaptation policies (19), highlighting a persistent gap in implementation and dissemination between national planning and frontline practice. The findings concurred with previous assertions that climate-sensitive plans to accelerate malaria elimination efforts in tropical regions and curricula for climate-adaptive strategies for healthcare workers, especially those in community roles, need to be developed (18,23–26).

The proposed mitigation and adaptation measures to combat the increasing malaria incidence due to climate change focused on community-level behavioural change strategies to reinforce existing malaria countermeasures, including vector control, reforestation, conservation farming and improving urban drainage. These are valid but do not yet reflect comprehensive health system resilience approaches required in a warming climate (2,13). Participants identified several contextual factors that undermine malaria control effectiveness, including resource diversion during the COVID-19 pandemic, the increased burden of other climate-sensitive infectious diseases such as cholera and arboviruses, and supply chain delays. A multifaceted approach could benefit countries struggling to eliminate malaria due to similar socio-economic practices within communities (31,32). Vector resistance, diagnostic inaccuracies and mobility patterns among farming and fishing communities were also noted as important barriers.

Despite observing climate-related shifts in malaria incidence, participants rarely mentioned the emerging role of malaria vaccines. This suggests an opportunity to integrate climate considerations into vaccine deployment strategies, especially given the quantitative component of this study, which shows that humidity, not rainfall or temperature, is the stronger climatic predictor of malaria trends in Zambia. Emerging evidence from the region suggests the potential role of malaria vaccines as a possible way to mitigate climate-driven disruptions in malaria control (29,33). These findings challenge longstanding assumptions that align vaccine timing primarily with rainfall patterns, reinforcing the need for climate-informed malaria vaccination strategies.

Taken together, these findings highlight a clear need for capacity-building initiatives that equip HCWs and CBVs with applied climate-health competencies, including interpreting climatic information, anticipating seasonal shifts, supporting community adaptation, and advocating for climate-resilient public health interventions (32,34). Climate change adaptation and mitigation strategies for malaria transmission should also include strengthening meteorological surveillance in health systems (8,15,35).

This study has several strengths. It draws on rich, diverse perspectives from HCWs across all provinces, capturing a wide range of professional experiences. The qualitative methods facilitated in-depth exploration of climate and health linkages, yielding insights with direct operational implications. Lessons outlined here of the frontline perspective may even be considered for mitigation strategies for other climate-sensitive infectious diseases, which are expected to rise (4,26,36,37).

There are limitations. Data were collected before the severe drought of 2024, the worst in Zambia’s recent history, as evidenced by the National Drought Emergency Response Plan (38), which may have further heightened the relevance of climate-health linkages. Nonetheless, the study findings remain valid, as both increased and decreased rainfall are recognised climate drivers of malaria transmission (19). The use of an established conceptual framework helped mitigate researcher bias and supported systematic interpretation of climate adaptation themes.

## Conclusion

Climate change is reshaping malaria transmission patterns in Zambia, and frontline health workers are already observing early impacts. Although HCWs and CBVs demonstrate strong knowledge of malaria control, significant gaps remain in their understanding of climate-related mechanisms and their application to climate-resilient strategies. Strengthening cross-sectoral collaboration, improving policy dissemination, and investing in climate-resilient health systems, including infrastructure, supply chains, and integrated climate-surveillance, are essential. Targeted training for HCWs and CBVs in climate-adaptive malaria control, together with community-level environmental stewardship, will be crucial to safeguard progress toward malaria elimination in a changing climate.

## Abbreviations

CBV: Community-Based Volunteer
EHO/T: Environmental Health Officer/Technician
IRS: Indoor Residual Spraying
ITN: Insecticide Treated Nets
ISDR: Integrated Disease Surveillance and Response
LLIN: Long Lasting Insecticide-treated Nets
NHCAP: National Health Climate Adaptation Plan
NMEC: National Malaria Control Centre
WHO: World Health Organization
ZNPHI: Zambia National Public Health Institute

## Data Availability

All the data is fully available, without restiction.

## Acknowledgements

This work is part of ongoing efforts by the Zambia National Public Health Institute and the National Malaria Elimination Centre to understand better bottlenecks towards achieving Malaria Elimination as a Country and in the Region. Many thanks to various team members at national and subnational levels, and partners who were directly and indirectly involved in this work.

## Institutional Review Board Statement

NHRA Approval (Ref No: NHRA000013/23/08/2023).

## Data availability statement

All relevant data are included in the paper.

## Author Contributions

**Conceptualisation:** Therese Shema Nzayisenga, Nyuma Mbewe, Kelvin Mwangilwa and Nathan Kapata

**Funding Acquisition:** Therese Shema Nzayisenga, Nyuma Mbewe, and Nathan Kapata

**Methodology and Data Curation**: Therese Shema Nzayisenga, Nyuma Mbewe, Kelvin Mwangilwa, Nathan Kapata, Allan Mayaba Mwiinde

**Supervision and Validation:** Nathan Kapata, Nyuma Mbewe, Allan Mayaba Mwiinde, Jonathan Mwanza

**Writing, reviewing and editing:** Nyuma Mbewe, Therese Shema Nzayisenga, Kelvin Mwangilwa, Jonathan Mwanza, Stephen Bwalya, Ignatius Banda, Cheepa Habeenzeu, Malambo Mutila, Paul Msanzya Zulu, Loveness Nikisi, Allan Mayaba Mwiinde, Nathan Kapata

